# Projected Population-Level Impact of Digital Return of Results for Cardiovascular–Kidney–Metabolic Screening at US Blood Donation Centers: A Monte Carlo Simulation Study

**DOI:** 10.64898/2026.09.01.26360806

**Authors:** Zehao Qian, Amit Khera, Sukh Makhnoon, Brian E. Chapman, Barbara Bryant, Merlyn Sayers, Frances Compton, Steven Eason, Chao Xing, Zahid Ahmad

## Abstract

**Background:** Cardiovascular–kidney–metabolic (CKM) syndrome affects nearly 90% of US adults, yet most individuals at early, modifiable stages remain unidentified outside clinical care. Blood donation centers offer a scalable, non-clinical venue for CKM screening, but the potential benefit of screening in this context remains unclear. We projected the population-level impact of effective digital return of results (ROR) to inform the design of a pragmatic trial.

**Methods:** We developed a Monte Carlo simulation (100,000 iterations) of the incident major adverse cardiovascular events (MACE), end-stage renal disease (ESRD), and type 2 diabetes (T2DM) preventable by ROR-prompted, guideline-concordant follow-up among donors in CKM Stages 1–2. The estimand counts only events averted by donors who act *because of* ROR; the intervention effect was modeled directly on strictly positive support, and action was translated into prevented events through a hazard-based cumulative-incidence difference that counts each donor at most once. We evaluated 18 design cells (donor volumes 300,000, 1 million, and 8 million/year; 5- and 10-year horizons; action-rate gains of +10, +20, and +30 percentage points [pp]) and, in a complementary two-arm simulation, the assurance (expected power) of detecting the effect in a single deployment.

**Results:** Under the primary +20 pp scenario, ROR at a single large blood center (300,000 donors/year) is projected to prevent a median of 2,201 events (95% uncertainty interval [UI], 1,099–4,364) over 10 years, scaling to 58,526 (29,154–116,769) at the national donor pool. All 18 design cells had strictly positive 95% lower bounds. The number needed to screen was 136 and the screening cost $2,045 per event prevented (at $15/donor), both invariant to donor volume. Impact scaled linearly with volume and effect size but sub-linearly with the horizon. Detection of the effect was effectively certain at gains of +20 pp or larger (assurance ≥99.6% in every cell and >99.9% in all but the smallest 5-year cell).

**Conclusions:** Even under the conservative scenario, digital CKM ROR at blood donation centers is projected to prevent hundreds to tens of thousands of incident cardiometabolic events at a screening cost per event well within accepted prevention benchmarks, providing prospective, quantitative justification for a pragmatic, randomized evaluation of digital ROR in non-clinical screening settings.

## 1 Introduction

Cardiovascular–kidney–metabolic (CKM) syndrome, the pathophysiologic continuum linking excess adiposity, metabolic dysfunction, chronic kidney disease, and cardiovascular risk, was formalized by the 2023 American Heart Association Presidential Advisory, which defined a five-stage classification (Stage 0, no risk factors, through Stage 4, established cardiovascular disease) and recommended screening for CKM risk factors across the life course.^1^ The burden is nearly universal: in nationally representative data, 89.4% of US adults meet criteria for CKM Stage 1 or higher, and roughly three-quarters of all US adults (74.9%) are at the early, modifiable Stages 1–2 rather than at advanced stages.^2^ Accompanying risk equations now embed CKM health in absolute cardiovascular risk prediction.^3^ Despite this mandate, most individuals at early stages remain unidentified outside clinical care.

Community blood donation centers are an underutilized venue for opportunistic CKM screening at population scale: US blood centers collect approximately 11.8 million red-cell units annually.^4^ Donation already involves phlebotomy and basic health screening, and donor eligibility criteria exclude most individuals with established cardiovascular disease or advanced kidney disease, concentrating the donor pool in CKM Stages 0–2. A recent pilot program at Carter BloodCare, a large community blood center in North Texas, demonstrated the feasibility of high-throughput cardiometabolic screening at donation: among 10,176 donors screened with non-fasting triglycerides, 39.2% had moderate and 2.4% severe hypertriglyceridemia, and most surveyed donors with severe elevations reported intent to seek care.^5^

A critical gap separates screening capacity from actionable result return. In clinical settings, individualized decision support and risk communication reliably improve knowledge, risk perception, and intent to act: a Cochrane review of 209 randomized trials of patient decision aids found large gains in knowledge and accuracy of risk perception with no adverse effect on decision regret,^6^ and a meta-analysis of 62 trials of cardiovascular risk communication found improved risk-factor trajectories and increased intention to begin preventive therapy.^7^ None of these strategies has been validated in the non-clinical blood-donation context, where donors lack clinician support. Before undertaking a large-scale pragmatic randomized trial of digital return of results (ROR) in this setting, it is important to quantify the potential public-health impact of successful implementation, together with the cost per event prevented and the probability that a deployment of realistic size could detect the effect.

Here we report a Monte Carlo simulation that projects the number of incident major adverse cardiovascular events (MACE), end-stage renal disease (ESRD), and type 2 diabetes (T2DM) preventable through effective digital CKM ROR at US blood donation centers, across a grid of donor volumes, time horizons, and intervention effect sizes. Two features of the specification are worth signposting. First, the estimand is *incremental*: only events averted by donors who act because of ROR are counted, so the assumed action-rate *gain* is the only behavioral input that must be defended. Second, action is translated into prevented events through a hazard-based cumulative-incidence difference rather than a linear rate-times-horizon product, so each donor is counted at most once and the marginal benefit declines appropriately as the at-risk pool is depleted. Together these choices yield uncertainty intervals with strictly positive lower bounds, without any floor or truncation, and a realistic, sub-linear response to the time horizon.

## 2 Methods

### 2.1 Model overview and estimand

For a design cell defined by annual donor volume *N*, time horizon *T*, and assumed action-rate gain *δ*, the model projects *Y*, the cumulative number of incident events prevented over *T* years that are *attributable to ROR*. This is an incremental quantity: it counts only events averted by the additional donors who take guideline-concordant action because of ROR, relative to the counterfactual without enhanced ROR. Events prevented by donors who would have acted anyway are excluded. The response is defined by

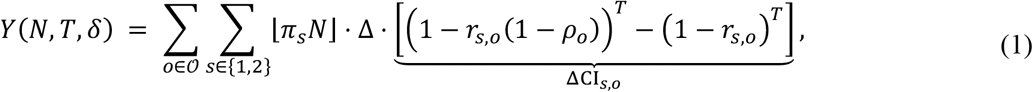

where *O* = {MACE, ESRD, T2DM} indexes outcomes, *s* indexes CKM stage (Stage 0 donors are screened but carry no modeled event risk; Stages 3–4 are excluded because donor eligibility criteria screen out established disease), π_*s*_ is the fraction of donors in stage *s*, Δ is the incremental action rate (the extra fraction of donors who act because of ROR), *r*_*s,o*_ is the stage- and outcome-specific annual event rate among non-actors, and *ρ*_*o*_ is the relative risk reduction achieved by acting. The per-actor factor ΔCI_*s,o*_ is the difference between the cumulative incidences at the untreated hazard *r*_*s,o*_ and at the treated hazard *r*_*s,o*_ (1 − *ρ*_*o*_), and is therefore the probability that acting averts the event within *T* years.

Equation (1) is the difference between the expected event counts of the counterfactual worlds with and without enhanced ROR (derivation in Supplementary Section S1); the baseline action rate does not appear in it, so the model specifies the effect Δ directly rather than reconstructing it as the difference between separately modeled baseline and post-intervention action rates. Because Δ is modeled on strictly positive support (below), consistent with the premise that returning a screening result does not reduce follow-up (a premise supported by the decision-aid and risk-communication trial literature^6,7^), every simulated draw contributes a non-negative benefit and no floor or truncation is applied anywhere in the pipeline.

### 2.2 Design grid

We evaluated three annual donor volumes: 300,000 (Carter BloodCare, from operational data), 1 million (a large regional center), and 8 million (the approximate national US donor pool, consistent in scale with the 11.8 million red-cell units collected annually^4^); two horizons (*T* = 5 and 10 years); and three action-rate gains (*δ* = 0.10, 0.20, 0.30, expressed as +10, +20, and +30 percentage points [pp]). The +20 pp gain is the primary scenario, with +10 and +30 pp as conservative and optimistic bounds; all three are design inputs to be tested in the planned trial, not empirical estimates, since no ROR trial in a blood-donor population exists. Each cell follows a single annual screening cohort for *T* years.

### 2.3 Stochastic specification and parameter sources

Uncertainty in every input was propagated by assigning each parameter a probability distribution and drawing *M* = 100,000 Monte Carlo samples, following standard practice for probabilistic analyses of decision models: Beta distributions for probability-type parameters, a Dirichlet distribution for the stage composition, and mean-preserving log-normal distributions for rates and for the strictly positive intervention effect.^8,9^ Table 1 lists each parameter, its distribution, and its source.

**Table 1.**
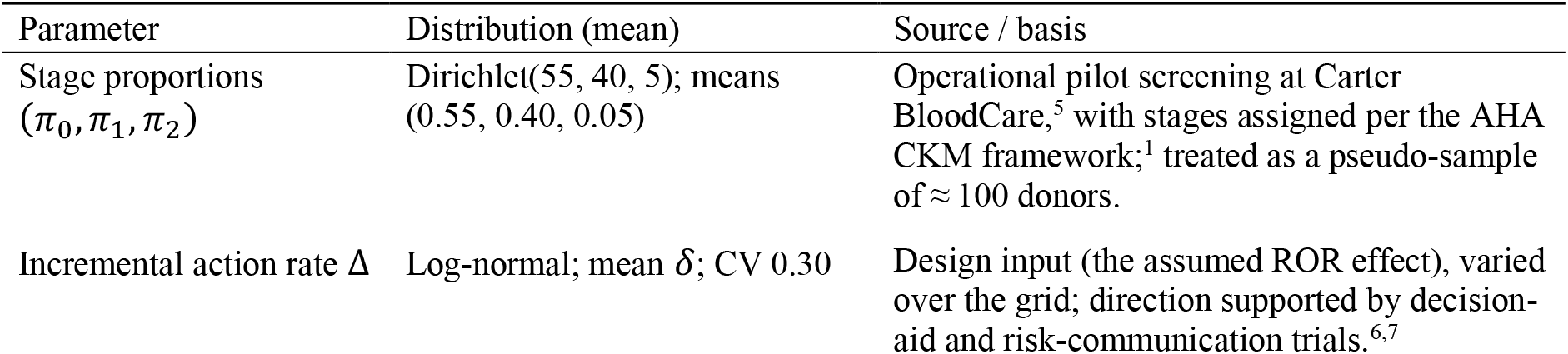
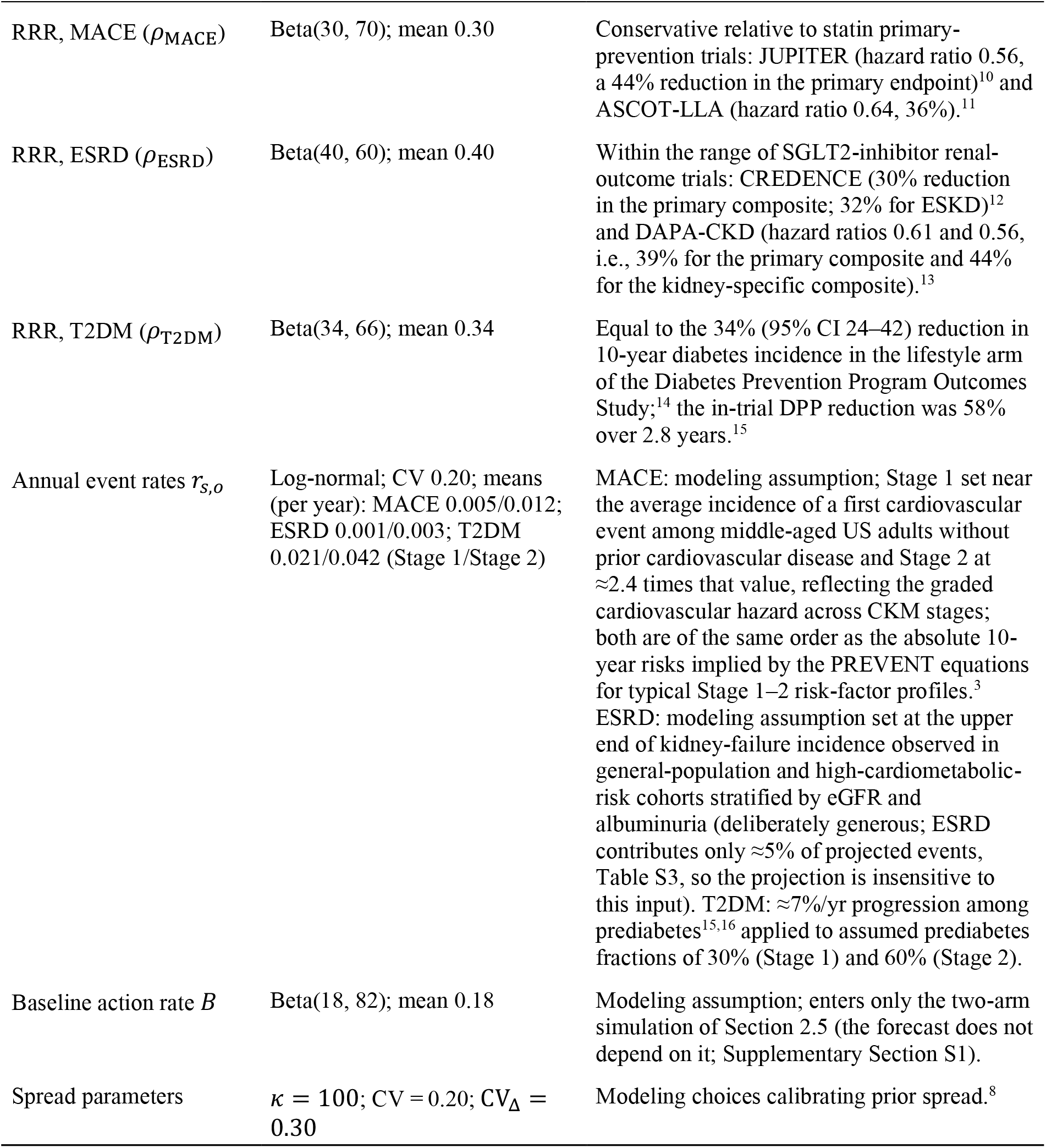
Model parameters, prior distributions, and sources. CV, coefficient of variation. The shared concentration *κ* = 100 acts as a pseudo–sample size, deliberately encoding the modest evidence base behind these inputs. Log-normal distributions use the mean-preserving parameterization, so each drawn quantity has exactly the stated mean.

| Parameter | Distribution (mean) | Source / basis |
| --- | --- | --- |
| Stage proportions<br>($\pi_0, \pi_1, \pi_2$) | Dirichlet(55, 40, 5); means<br>(0.55, 0.40, 0.05) | Operational pilot screening at Carter BloodCare,<sup>5</sup> with stages assigned per the AHA CKM framework;<sup>1</sup> treated as a pseudo-sample of $\approx 100$ donors. |
| Incremental action rate $\Delta$ | Log-normal; mean $\delta$; CV 0.30 | Design input (the assumed ROR effect), varied over the grid; direction supported by decision-aid and risk-communication trials.<sup>6,7</sup> |
| RRR, MACE ($\rho_{\text{MACE}}$) | Beta(30, 70); mean 0.30 | Conservative relative to statin primary-prevention trials: JUPITER (hazard ratio 0.56, a 44% reduction in the primary endpoint)<sup>10</sup> and ASCOT-LLA (hazard ratio 0.64, 36%).<sup>11</sup> |
| RRR, ESRD ($\rho_{\text{ESRD}}$) | Beta(40, 60); mean 0.40 | Within the range of SGLT2-inhibitor renal-outcome trials: CREDENCE (30% reduction in the primary composite; 32% for ESKD)<sup>12</sup> and DAPA-CKD (hazard ratios 0.61 and 0.56, i.e., 39% for the primary composite and 44% for the kidney-specific composite).<sup>13</sup> |
| RRR, T2DM ($\rho_{\text{T2DM}}$) | Beta(34, 66); mean 0.34 | Equal to the 34% (95% CI 24–42) reduction in 10-year diabetes incidence in the lifestyle arm of the Diabetes Prevention Program Outcomes Study;<sup>14</sup> the in-trial DPP reduction was 58% over 2.8 years.<sup>15</sup> |
| Annual event rates $r_{s,o}$ | Log-normal; CV 0.20; means (per year): MACE 0.005/0.012; ESRD 0.001/0.003; T2DM 0.021/0.042 (Stage 1/Stage 2) | MACE: modeling assumption; Stage 1 set near the average incidence of a first cardiovascular event among middle-aged US adults without prior cardiovascular disease and Stage 2 at $\approx 2.4$ times that value, reflecting the graded cardiovascular hazard across CKM stages; both are of the same order as the absolute 10-year risks implied by the PREVENT equations for typical Stage 1–2 risk-factor profiles.<sup>3</sup> ESRD: modeling assumption set at the upper end of kidney-failure incidence observed in general-population and high-cardiometabolic-risk cohorts stratified by eGFR and albuminuria (deliberately generous; ESRD contributes only $\approx 5\%$ of projected events, Table S3, so the projection is insensitive to this input). T2DM: $\approx 7\%/yr$ progression among prediabetes<sup>15,16</sup> applied to assumed prediabetes fractions of 30% (Stage 1) and 60% (Stage 2). |
| Baseline action rate $B$ | Beta(18, 82); mean 0.18 | Modeling assumption; enters only the two-arm simulation of Section 2.5 (the forecast does not depend on it; Supplementary Section S1). |
| Spread parameters | $\kappa = 100$; CV = 0.20; $CV_{\Delta} = 0.30$ | Modeling choices calibrating prior spread.<sup>8</sup> |

Two conventions matter for interpretation. First, all log-normal parameterizations are mean-preserving (µ = ln *m* − ½ σ^2^, σ^2^ = ln(1 + CV^2^)), so E[Δ] = *δ* and E[*r*_*s,o*_] = *m*_*s,o*_ exactly. Second, the stage proportions, risk reductions, and event rates are drawn once and reused across all 18 design cells (common random numbers), so cells differ only by design and not by simulation noise; only Δ is redrawn per cell, because its mean is the cell’s design input. Because the input distributions encode prior evidence and are not updated against outcome data, the reported 2.5th–97.5th percentile ranges are prior-predictive uncertainty intervals (UIs) rather than posterior credible intervals. Figure 1 summarizes the simulation pipeline; pseudocode is given in Supplementary Section S2 (Algorithm 1).

**Figure 1.**
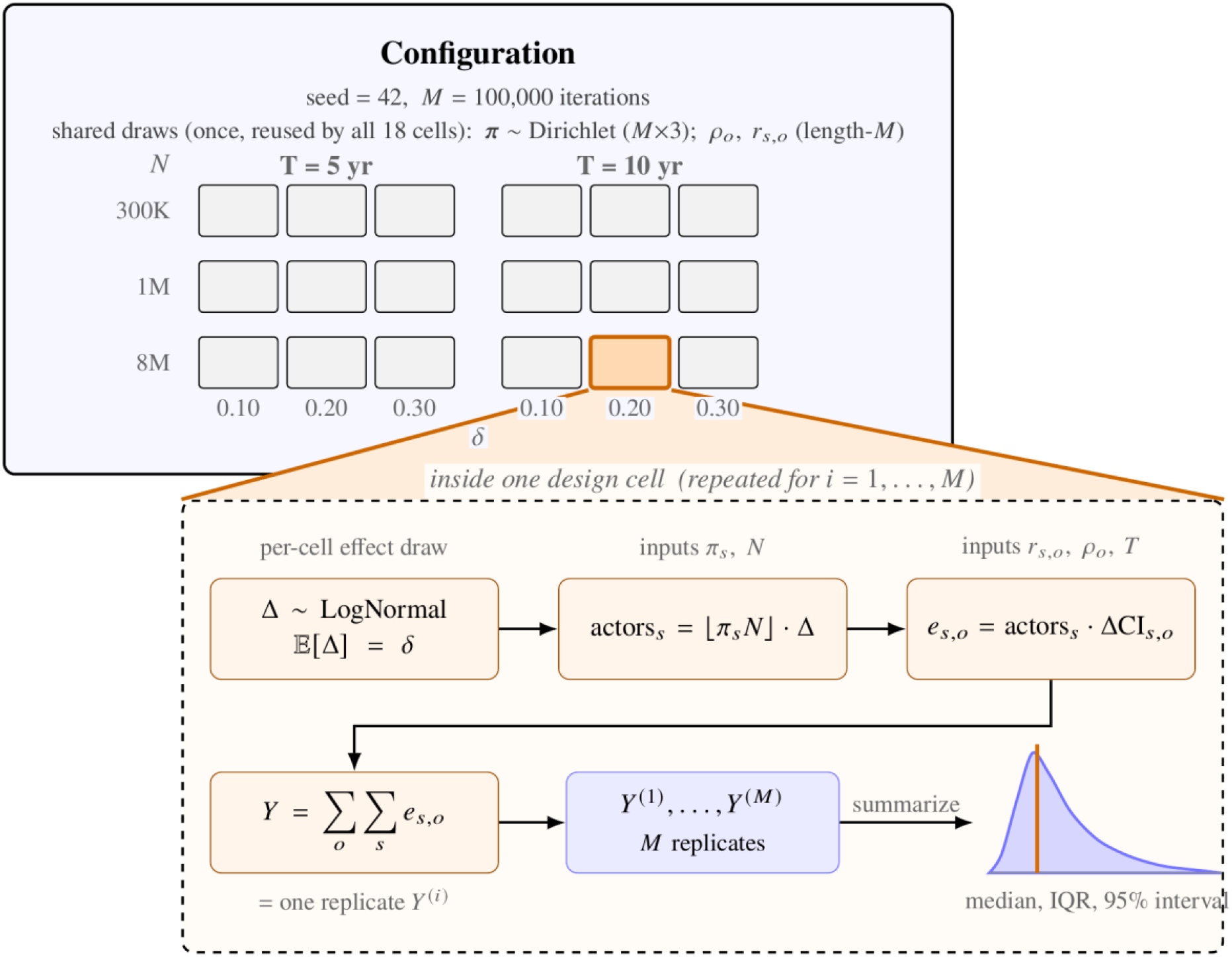
Overview of the simulation. The *Configuration* box holds the fixed inputs and the once-drawn shared parameters, together with the 18 design cells indexed by donor volume *N*, horizon *T*, and action-rate gain *δ*. Magnifying any single cell shows the per-cell computation: one Monte Carlo replicate *Y*^(*i*)^ is produced by drawing the incremental action rate Δ, forming the incremental actors, converting them into events prevented through the cumulative-incidence difference ΔCI_*s,o*_, and summing over stages and outcomes. Repeating this for *i* = 1, …, *M* yields the length-*M* sample, whose distribution is summarized by the median, IQR, and 95% uncertainty interval.

### 2.4 Event translation

Because an incident event can occur at most once per person, per-person benefit is a cumulative-incidence difference, not an annual rate multiplied by the horizon. Under a constant annual hazard *r*, the probability of at least one event within *T* years is 1 − (1 − *r*)^*T*^; the per-actor probability that acting averts the event is the difference of the treated and untreated cumulative incidences, ΔCI_*s,o*_ in equation (1). This translation counts each donor at most once and induces the model’s sub-linear horizon response: the factor saturates as *T* grows, fastest for the highest-incidence outcome (Supplementary Table S1).

### 2.5 Two-arm simulation and assurance

To characterize detectability, we additionally simulated a parallel-group evaluation: a control cohort of *N* donors with action rate *B* and an ROR cohort of *N* donors with action rate *B* + Δ, sharing each Monte Carlo replication’s parameter draws (common random numbers) but realized independently. Within each arm, actor counts and realized events are binomial at the appropriate cumulative-incidence probabilities. The between-arm difference in total realized events, *D*_*E*_, was tested with a normal-approximation score statistic for the difference of two Poisson counts, 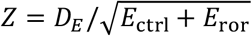, one-sided α = 0.05. Because the arm totals are sums of binomials, whose variance is smaller than the Poisson value, the test is slightly conservative; the discrepancy is largest (≈10–15% of the standard deviation) for T2DM. Averaging the indicator of significance over the 100,000 replications, each of which draws fresh parameters and a fresh realization, yields the *assurance*, or expected power: the unconditional probability that a single deployment returns a significant result, i.e., classical power averaged over the prior on the effect and the other parameters.^17^ By construction the expected between-arm difference equals the forecast estimand, so the two analyses are consistent, and the two-arm simulation adds specifically the sampling-uncertainty statement the forecast cannot provide. In addition to the three primary gains, we evaluated the assurance over a finer grid of action-rate gains (+2 to +30 pp in 2-pp steps) at every volume and horizon, to locate the smallest gain each deployment size can reliably detect.

### 2.6 Implementation

The simulation was implemented in R (version 4.6.1) with fixed seed (42) and *M* = 100,000 iterations per design cell; with *M* = 100,000, Monte Carlo error in reported medians is well under 1%. Reported summaries are the median, interquartile range (IQR), and 95% UI (2.5th–97.5th percentiles). Analysis code and all generated results are provided with this manuscript (Data availability). This study used only published aggregate data and simulation; no individual-level human data were involved.

## 3 Results

### 3.1 Projected events prevented

Table 2 and Figure 2 summarize the total projected events prevented in all 18 design cells; outcome-specific results appear in Supplementary Tables S2–S3. Under the primary scenario (+20 pp), effective digital CKM ROR at a single large blood center (300,000 donors/year) is projected to prevent a median of 2,201 events (IQR 1,736–2,791; 95% UI 1,099–4,364) over 10 years, rising to 7,326 (95% UI 3,658–14,579) at a regional center and 58,526 (95% UI 29,154–116,769) at national scale. Every one of the 18 cells has a strictly positive 95% lower bound (minimum across cells: 295 events, in the smallest and most conservative cell); because the effect has positive support by construction, no probability mass accumulates at zero and the intervals require no floor, truncation, or caveat.

**Table 2.** Projected ROR-attributable events prevented (all outcomes combined) for a single annual screening cohort, by donor volume, horizon, and assumed action-rate gain. Cells give the median (95% uncertainty interval) over 100,000 Monte Carlo iterations. Interquartile ranges and outcome-specific results are given in Supplementary Tables S2–S3.

| Effect size | Horizon | 300,000/yr | 1,000,000/yr | 8,000,000/yr |
| --- | --- | --- | --- | --- |
| +10 pp (conservative) | 5 yr | 597 (295–1,201) | 1,995 (990–4,019) | 15,935 (7,894–32,076) |
|  | 10 yr | 1,099 (548–2,190) | 3,669 (1,817–7,330) | 29,351 (14,644–58,188) |
| +20 pp (primary) | 5 yr | 1,197 (590–2,410) | 3,989 (1,967–8,002) | 31,941 (15,798–64,251) |
|  | 10 yr | 2,201 (1,099–4,364) | 7,326 (3,658–14,579) | 58,526 (29,154–116,769) |
| +30 pp (optimistic) | 5 yr | 1,794 (885–3,605) | 5,986 (2,958–12,101) | 47,775 (23,684–96,315) |
|  | 10 yr | 3,294 (1,643–6,575) | 10,979 (5,490–21,822) | 88,030 (43,609–175,157) |

**Figure 2.**
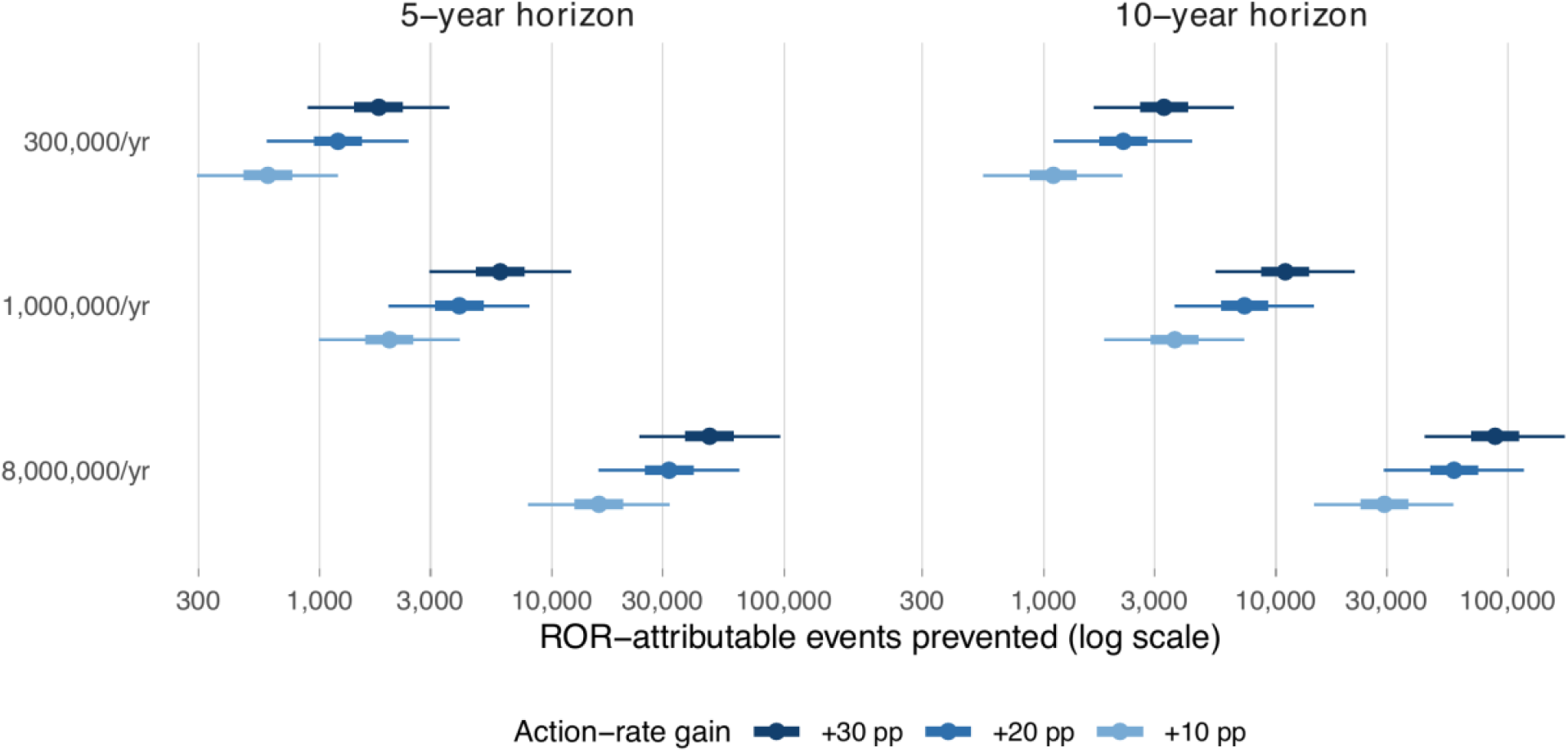
Projected ROR-attributable events prevented across the design grid. Points are medians, thick bars interquartile ranges, and thin bars 95% uncertainty intervals over 100,000 Monte Carlo iterations, shown on a logarithmic scale by annual donor volume and action-rate gain, for the 5-year (left) and 10-year (right) horizons.

The projection scales in exact proportion to donor volume and to the assumed action-rate gain, both of which enter equation (1) multiplicatively. The horizon does not: doubling the horizon from 5 to 10 years multiplies the projected impact by 1.84 rather than 2, because donors who have already experienced an event are no longer at risk. The shortfall is concentrated in the highest-incidence outcome: evaluated at prior means, the per-actor benefit ratio between horizons is 1.90–1.99 for MACE and ESRD but 1.68 for Stage 2 T2DM (Supplementary Table S1). Incident T2DM contributes 75.4% of the total in the primary 10-year cell, MACE 19.0%, and ESRD 5.6%; the ordering follows the epidemiology, since T2DM incidence in Stages 1–2 is an order of magnitude higher than that of MACE or ESRD. The T2DM share declines slightly with the horizon (76.9% at 5 years) as its cumulative incidence saturates first.

### 3.2 Planning quantities

Table 3 reports the number needed to screen (NNS) and the screening cost per event prevented, assuming $15 per donor screened (midpoint of the $10–$20 operational range). For the primary scenario at the 10-year horizon, 136 donors must be screened per event prevented, at a screening cost of $2,045 per event. Because the projected impact scales linearly with volume, both quantities are invariant to donor volume: scaling from a single center to the national pool multiplies the screening budget and the events prevented by the same factor. The genuine variation is across effect size and horizon: both quantities are inversely proportional to the gain *δ*, and are reduced by somewhat less than half when the horizon doubles. Both quantities are defined against a single annual cohort and its *T*-year event yield, and the cost base includes the screening test only, not downstream care; the figure is a screening cost per event, not a cost-effectiveness ratio.

**Table 3.**
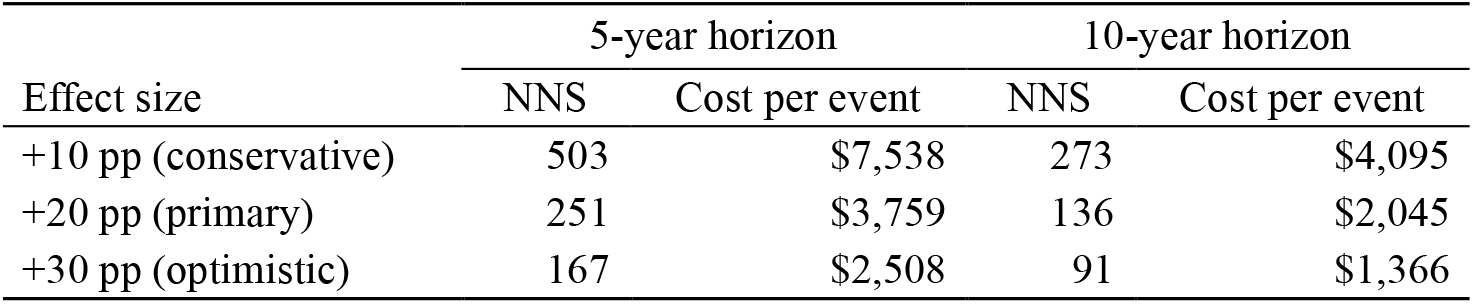
Number needed to screen (NNS, donors screened per event prevented) and screening cost per event prevented, by action-rate gain and horizon, at $15 per donor screened. Both quantities are invariant to donor volume (medians differ across volumes only by Monte Carlo noise and rounding); they are defined for a single annual cohort followed over the stated horizon, and the cost base is the screening test only.

| Effect size | 5-year horizon |  | 10-year horizon |  |
| --- | --- | --- | --- | --- |
|  | NNS | Cost per event | NNS | Cost per event |
| +10 pp (conservative) | 503 | \$7,538 | 273 | \$4,095 |
| +20 pp (primary) | 251 | \$3,759 | 136 | \$2,045 |
| +30 pp (optimistic) | 167 | \$2,508 | 91 | \$1,366 |

### 3.3 Detectability of the effect in a single deployment

Table 4 reports the assurance, defined as the fraction of 100,000 simulated two-arm deployments in which the one-sided test reached *p* < 0.05, for every design cell; full arm-level event counts and median *p*-values appear in Supplementary Table S4. In the reference cell the two-arm simulation reproduces the forecast, with a median between-arm difference of 2,210 events against the forecast median of 2,201. Figure 3 contrasts the two arms for this cell. Marginally, the arms’ realized event totals overlap heavily, because parameter uncertainty moves both cohorts together and the ROR arm’s distribution is therefore only shifted modestly relative to the control arm’s (median 32,441 versus 34,782 events; Figure 3A). Within replications, however, the two arms share their parameter draws, and the between-arm difference isolates the intervention: *D*_*E*_ was positive in every one of the 100,000 simulated trials, with 95% of differences between 997 and 4,430 events (Figure 3B). At gains of +20 pp and above, detection is effectively certain (assurance 99.6% for a 5-year deployment at a 300,000-donor center and >99.9% in every other cell). Assurance falls below 99% only under the conservative +10 pp gain at the smallest volume: 88.4% for a 5-year deployment at a 300,000-donor center, rising to 96.7% over 10 years, and already 99.3% at the regional volume over 5 years. The median *p*-value is below 10^−3^ in every cell. A pragmatic evaluation of digital CKM ROR at realistic scale is therefore well-powered essentially throughout the design space, with the single weakest combination (a small center, the conservative gain, and the short horizon) still detecting the effect in roughly nine deployments in ten.

**Table 4.** Assurance (expected power, %) of a two-arm deployment: the proportion of 100,000 simulated trials in which the one-sided difference-of-Poisson-counts test reached *p* < 0.05, averaging over both parameter uncertainty and sampling variability. Arms are equal-sized cohorts at the stated annual volume.

| Donor volume | Horizon | +10 pp | +20 pp | +30 pp |
| --- | --- | --- | --- | --- |
| 300,000/yr | 5 yr | 88.3 | 99.6 | >99.9 |
|  | 10 yr | 96.7 | >99.9 | >99.9 |
| 1,000,000/yr | 5 yr | 99.3 | >99.9 | >99.9 |
|  | 10 yr | 99.9 | >99.9 | >99.9 |
| 8,000,000/yr | 5 yr | >99.9 | >99.9 | >99.9 |
|  | 10 yr | >99.9 | >99.9 | >99.9 |

**Figure 3.**
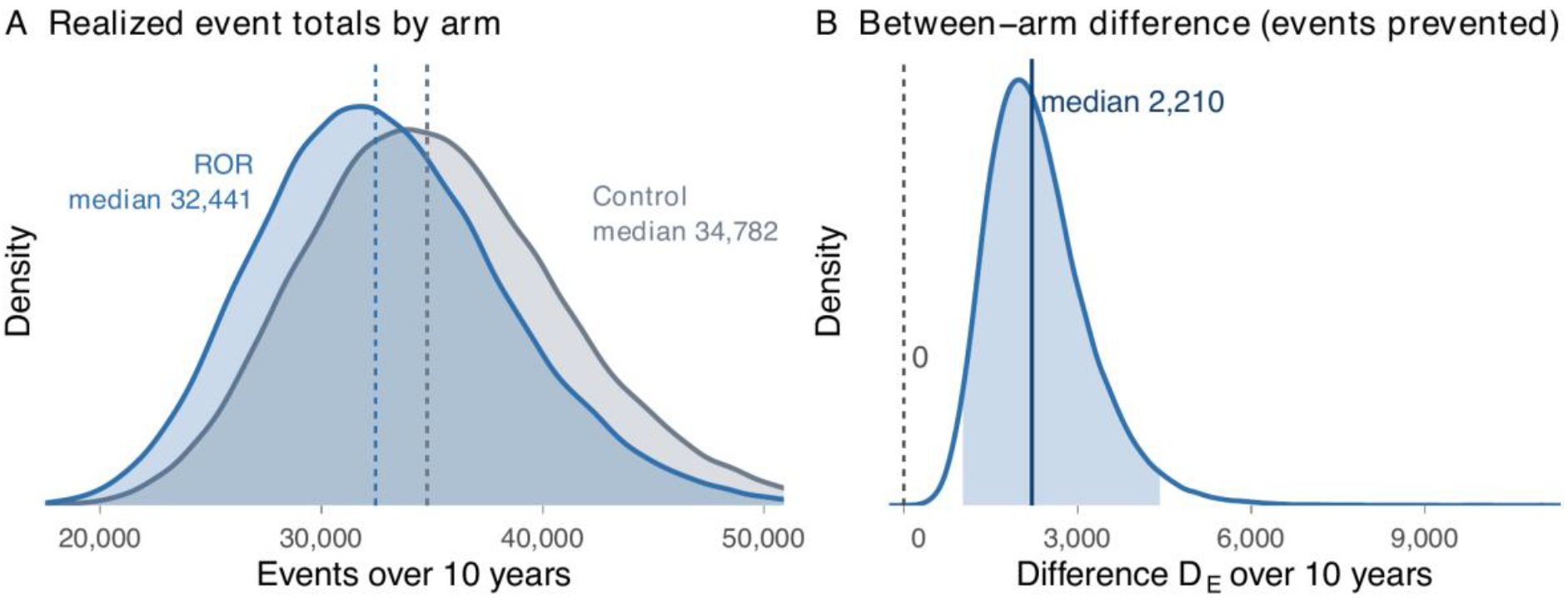
Control versus ROR arm in the simulated two-arm evaluation, for the reference cell (300,000 donors per arm, 10-year horizon, +20 pp gain; 100,000 simulated trials). (A) Distributions of the realized 10-year event totals in each arm (dashed lines, medians). The heavy overlap reflects parameter uncertainty, which moves both cohorts together. (B) Distribution of the between-arm difference *D*_*E*_ (events prevented), which cancels the shared parameter draws and isolates the intervention: the difference is positive in every replication. The shaded region is the 95% uncertainty interval (997–4,430); the solid line marks the median (2,210).

The finer effect-size grid locates the smallest gain each deployment size can reliably detect, and the detection threshold falls steeply with scale: 90% assurance requires a gain of roughly +12 pp over five years (+8 pp over ten) at a 300,000-donor center, +6 pp at 1 million donors, and only +2 to +4 pp at the national scale. The proposed evaluation therefore retains high assurance even for effects well below the conservative scenario.

## 4 Discussion

This simulation projects that effective digital return of CKM screening results at US blood donation centers would prevent a substantial number of incident cardiometabolic events at a screening cost of about $2,000 per event prevented: a median of roughly 2,200 over 10 years for a single large center under the primary assumption of a +20 pp gain in guideline-concordant follow-up, and roughly 59,000 at the scale of the national donor pool. The uncertainty intervals around these projections exclude zero in every scenario examined, including the most conservative, and a pragmatic two-arm evaluation of the strategy would be adequately powered at every donor volume considered.

Two specification choices drive the behavior of these estimates, and both were made for identifiable methodological reasons. First, the intervention effect is modeled directly as a single strictly positive quantity rather than reconstructed as the difference of two independently uncertain action rates. This yields uncertainty intervals whose strictly positive lower bounds arise from the model structure itself, not from flooring, truncation, or post hoc reinterpretation. It also concentrates the model’s key behavioral assumption in a single quantity, the gain *δ*, which is transparent about what the planned trial must actually measure. Second, the hazard-based cumulative-incidence translation counts each donor at most once, which lowers the projection relative to a linear rate-times-horizon calculation (most for the dominant T2DM outcome) and produces the realistic sub-linear response to the horizon observed in the results. Both choices make the present projections more conservative than a naive specification, not less.

The projections compare favorably with accepted prevention benchmarks. The NNS of 136 (primary scenario, 10 years) is of the same order as established screening programs for cardiovascular risk, and the screening cost per event prevented of about $2,000 sits well below typical willingness-to-pay thresholds for averting a major cardiovascular or renal event or an incident diabetes case, even though it omits, by design, downstream treatment costs and any benefit accruing beyond the modeled first events. The two-arm analysis adds the planning quantity a projection alone cannot supply: because parameters are drawn from their priors rather than fixed at a single alternative, the reported detection probability is an assurance in the sense of O’Hagan et al.,^17^ the appropriate power notion for a prior-averaged design analysis. That assurance is at least 99.6% at the primary effect size in every cell indicates that the proposed evaluation is not gated on optimistic assumptions about the effect.

### 4.1 Limitations

Several limitations warrant emphasis. First, the three outcomes are sampled independently, whereas MACE, ESRD, and T2DM share metabolic determinants and are positively correlated within individuals. Because expectations add regardless of dependence, the central projections are essentially unaffected, but the total’s uncertainty intervals are likely somewhat too narrow and the reported assurance correspondingly slightly optimistic; coupling the outcomes through a shared frailty or copula is the natural refinement. Second, each relative risk reduction is applied as a constant proportional hazard reduction across the whole horizon. Real intervention effects attenuate as adherence decays, so the constant *ρ*_*o*_ should be read as a horizon-averaged summary; the 10-year T2DM value is anchored to a follow-up of that same length,^14^ but if true effect trajectories decline, the model places too much benefit in later years. Third, competing risks across outcomes are not modeled; the cumulative-incidence translation removes within-outcome depletion, which is the larger error, but a donor removed by one event remains at risk of the others in the model. Fourth, the MACE and ESRD annual event rates are order-of-magnitude assumptions set at the scale of published risk equations and eGFR/albuminuria risk strata^3,18^ rather than direct estimates from a staged donor population (the ESRD values in particular are generous relative to general-population incidence, but ESRD carries ≈5% of the projected total), and the T2DM rates inherit an assumed prediabetes fraction by stage; the stage distribution itself derives from a single pilot program.^5^ Fifth, the effect spread (CV_Δ_ = 0.30) is a modeling choice, since no ROR trial in a blood-donor population exists to calibrate it; interval widths are directly sensitive to it. Finally, the relative risk reductions derive from trial populations receiving active interventions under trial conditions; real-world attenuation of adherence after initiation would reduce the projected benefit, and this is the dominant downside risk to the central estimates.

### 4.2 Conclusions

Under deliberately structured and largely conservative assumptions, digital CKM ROR at blood donation centers is projected to prevent thousands of incident cardiometabolic events at a single large blood center and tens of thousands nationally over a 10-year horizon, at a screening cost per event well within accepted prevention benchmarks, and with near-certain detectability in a pragmatic two-arm evaluation at realistic scale. These projections provide prospective, quantitative justification for a randomized evaluation of digital ROR strategies in non-clinical screening settings, and the modeling framework, whose behavioral input reduces to the single action-rate gain that such a trial would directly measure, is designed to be updated as empirical data accrue.

## Supporting information

Simulation Scripts

Simulation Results

## Data Availability

This study is a Monte Carlo simulation and used no individual-level data. All model inputs are aggregate estimates from previously published sources cited in the manuscript (Table 1 and reference list). The complete analysis code (R scripts implementing the forecast model, the two-arm simulation, and all tables and figures) and all generated simulation results are openly available at https://github.com/dukechain2333/blood_donor_ROR_simulation and permit exact reproduction of every reported number (fixed seed 42, 100,000 iterations).

## Ethics statement

This study is a computer simulation using only published aggregate estimates and operational planning figures; it involved no human participants, no individual-level data, and no identifiable information, and did not require institutional review board approval.

## Data and code availability

All analysis code (R scripts implementing the forecast model, the two-arm simulation, and all tables and figures) and the complete generated results accompany this manuscript and permit exact reproduction of every reported number (fixed seed, 100,000 iterations). No individual-level data were used.

## Supplementary Material

### S1 Invariance of the projection to the baseline action rate

Fix a stage *s* and outcome *o*, write *n*_*s*_ = ⌊π_*s*_*N*⌋ for the stage head-count, and define the per-person *T*-year cumulative incidences without and with action,

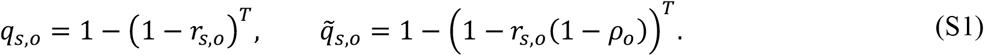

In the counterfactual world without enhanced ROR a fraction *B* of donors act, so the expected events are

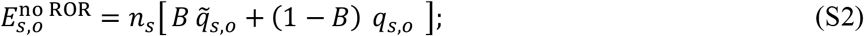

with ROR the acting fraction rises to *B* + Δ, giving

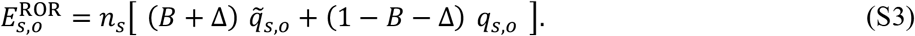

Their difference is the ROR-attributable increment:

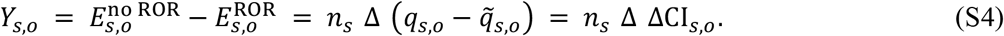

Every term involving *B* cancels exactly, for every Monte Carlo draw, so the projection *Y* = ∑_*o*_ ∑_*s*_ *Y*_*s,o*_ is invariant to the baseline action rate. Two consequences follow. First, the baseline need not be specified for the forecast at all; a value is required only by the two-arm simulation, which models realized (not incremental) event counts. Second, modeling the effect Δ directly, rather than as the difference of independently drawn baseline and post-intervention rates, removes nuisance baseline-location variance from the effect and, with log-normal (positive) support, guarantees a non-negative modeled benefit in every iteration without any floor or truncation. In the implementation the baseline is drawn after every quantity the forecast consumes, so changing its assumed mean cannot perturb the forecast’s random-number stream: the forecast is numerically, not merely statistically, invariant to it.

### S2 Stochastic specification and algorithm

The generative model for one Monte Carlo iteration is

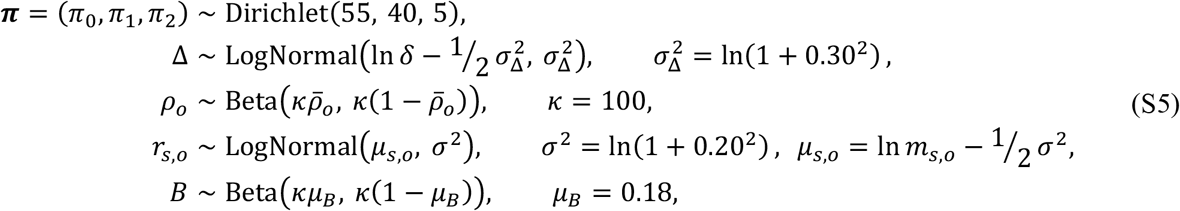

with prior mean risk reductions 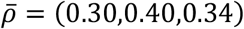 for (MACE, ESRD, T2DM) and prior mean annual rates *m*_*s,o*_ as in Table 1 of the main text. Both log-normal parameterizations are mean-preserving. The Beta and Dirichlet priors are written in mean–concentration form; the shared concentration *κ* = 100 acts as a pseudo–sample size of roughly one hundred observations per input.

The complete forecast procedure is given in Algorithm 1.

#### Algorithm 1

CKM ROR Monte Carlo Forecast

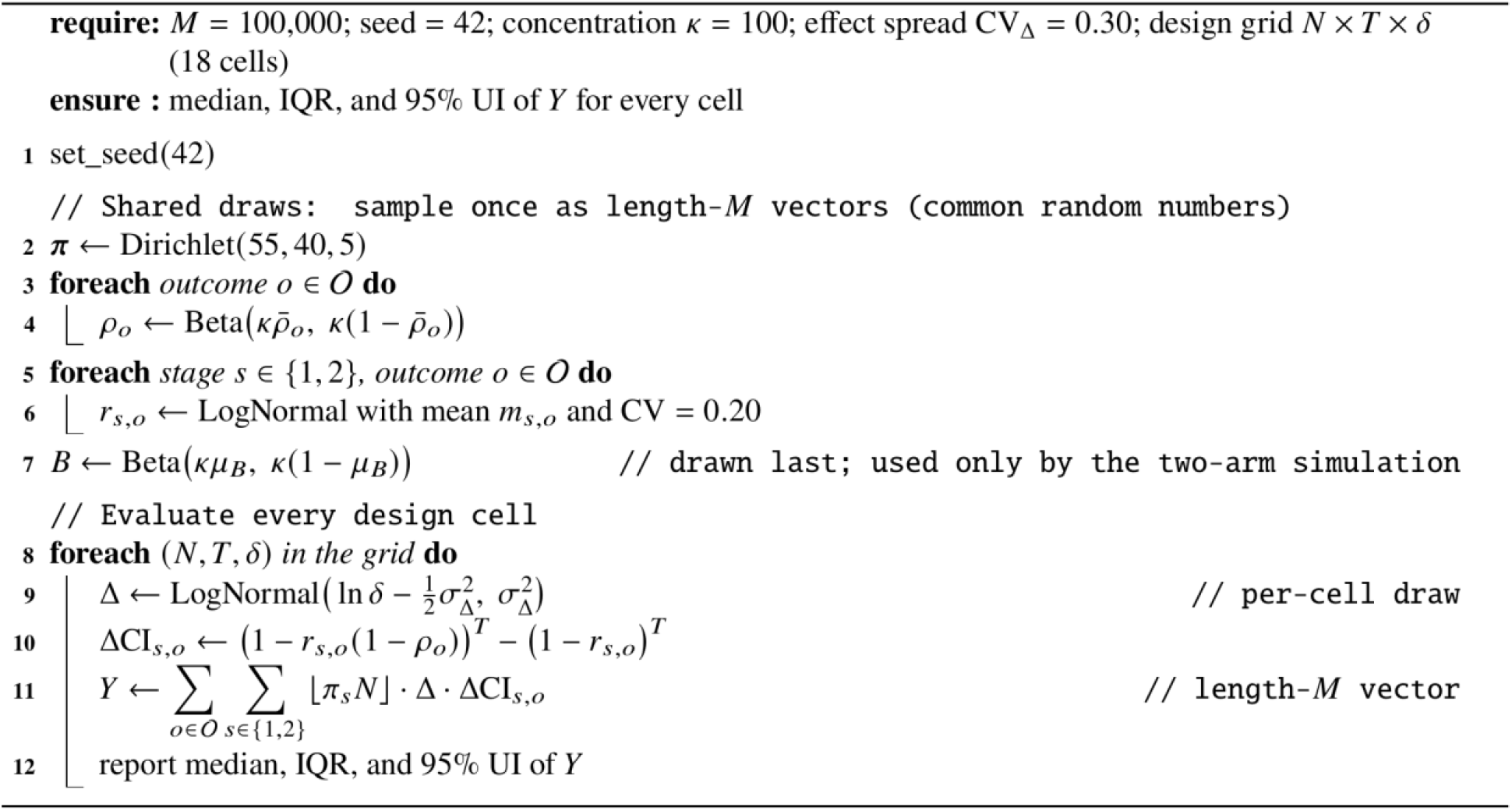

For the two-arm simulation (main text, Section 2.5), each replication additionally realizes actor counts 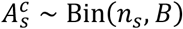 and 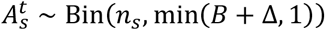 and realized events as binomial draws at the cumulative-incidence probabilities 1 − (1 − *r*_*s,o*_)^T^ (non-actors) and 1 − (1 − *r*_*s,o*_ (1 − *ρ*_*o*_))^T^ (non-actors) and 1 − (1 − *r*_*s,o*_ (1 − *ρ*_*o*_))^T^ (actors), summed over stages and outcomes within each arm.

### S3 Per-actor benefit and its saturation over the horizon

Table S1 evaluates the per-actor cumulative-incidence difference ΔCI_*s,o*_ at the prior mean rates and risk reductions for the two evaluated horizons. A translation linear in the horizon would give a 10-year/5-year ratio of exactly 2 in every row; the shortfall measures depletion of the at-risk pool, and is material only for the highest-incidence outcome (Stage 2 T2DM, ratio 1.68), whereas the MACE and ESRD rows remain nearly linear in the horizon.

**Table S1.** Per-actor probability that an event is averted, ΔCI_*s,o*_ = (1 − *r*(1 − *ρ*))^*T*^ − (1 − *r*)^*T*^, evaluated at prior mean annual rate *r* and mean relative risk reduction *ρ*, by horizon.

| Outcome, stage | $r$ | $\rho$ | $\Delta \text{CI} (T = 5)$ | $\Delta \text{CI} (T = 10)$ | 10-yr / 5-yr |
| --- | --- | --- | --- | --- | --- |
| MACE, Stage 1 | 0.005 | 0.30 | 0.0074 | 0.0144 | 1.96 |
| MACE, Stage 2 | 0.012 | 0.30 | 0.0173 | 0.0328 | 1.90 |
| ESRD, Stage 1 | 0.001 | 0.40 | 0.0020 | 0.0040 | 1.99 |
| ESRD, Stage 2 | 0.003 | 0.40 | 0.0059 | 0.0117 | 1.98 |
| T2DM, Stage 1 | 0.021 | 0.34 | 0.0333 | 0.0610 | 1.83 |
| T2DM, Stage 2 | 0.042 | 0.34 | 0.0620 | 0.1038 | 1.68 |

### S4 Full simulation results

**Table S2.**
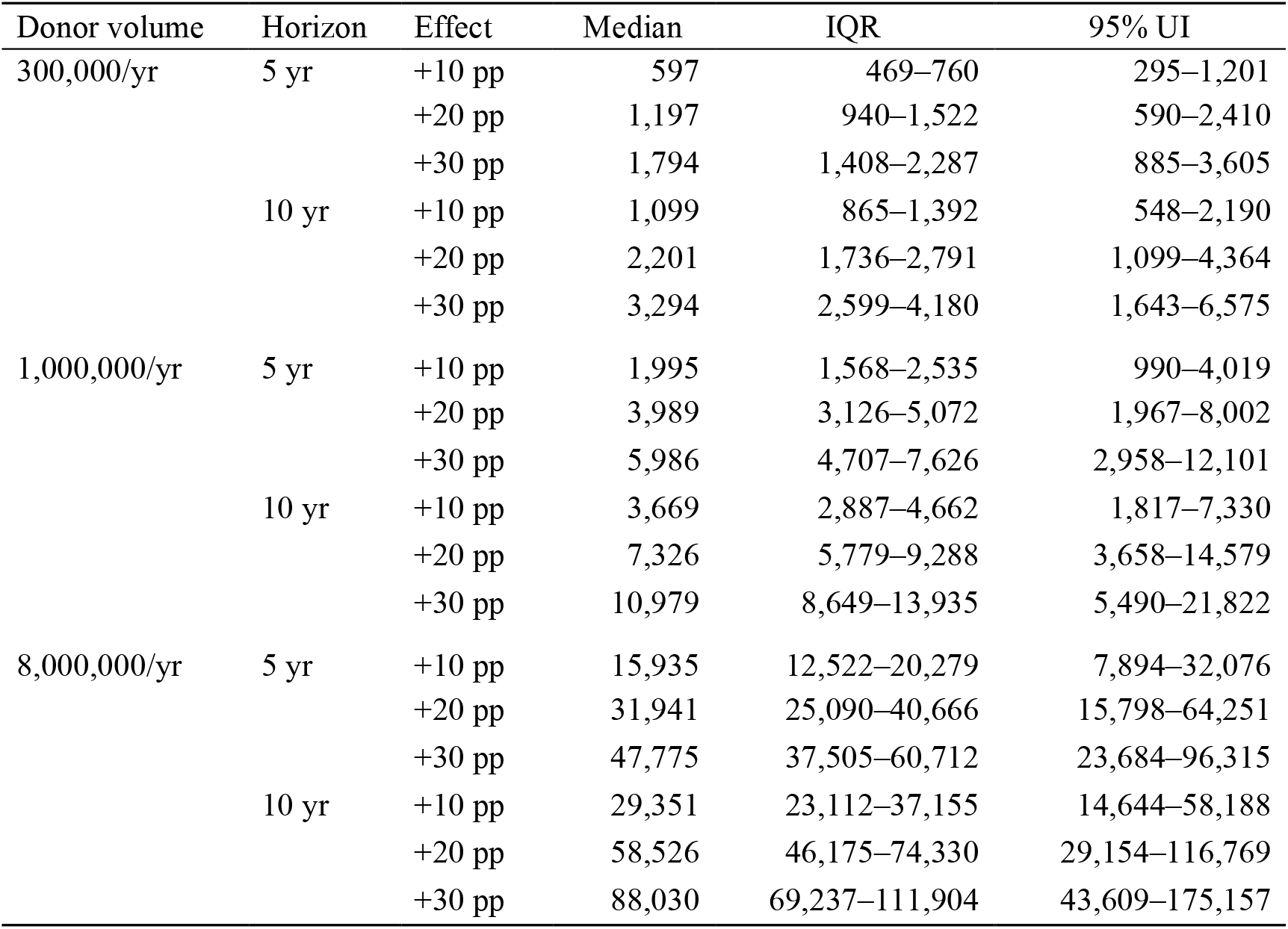
Total ROR-attributable events prevented in all 18 design cells: median, interquartile range (IQR), and 95% uncertainty interval (UI) over 100,000 Monte Carlo iterations.

**Table S3.** Outcome-specific projected events prevented under the primary scenario (+20 pp): median (share of the total, computed on means), with the combined total. Shares are invariant to donor volume up to rounding; they shift slightly with the horizon as the T2DM cumulative incidence saturates.

| Donor volume | Horizon | MACE | ESRD | T2DM | Total |
| --- | --- | --- | --- | --- | --- |
| 300,000/yr | 5 yr | 212 (17.9%) | 61 (5.1%) | 915 (76.9%) | 1,197 |
|  | 10 yr | 414 (19.0%) | 121 (5.6%) | 1,650 (75.4%) | 2,201 |
| 1,000,000/yr | 5 yr | 707 (17.9%) | 203 (5.1%) | 3,044 (76.9%) | 3,989 |
|  | 10 yr | 1,377 (19.0%) | 405 (5.6%) | 5,489 (75.4%) | 7,326 |
| 8,000,000/yr | 5 yr | 5,675 (17.9%) | 1,631 (5.1%) | 24,428 (76.9%) | 31,941 |
|  | 10 yr | 11,022 (19.0%) | 3,239 (5.6%) | 43,850 (75.4%) | 58,526 |

**Table S4.**
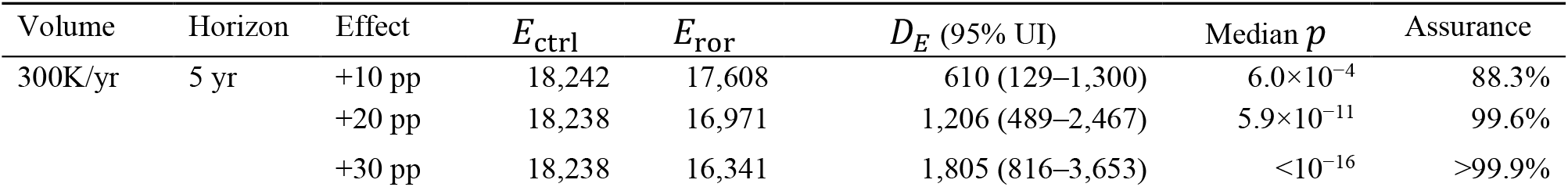
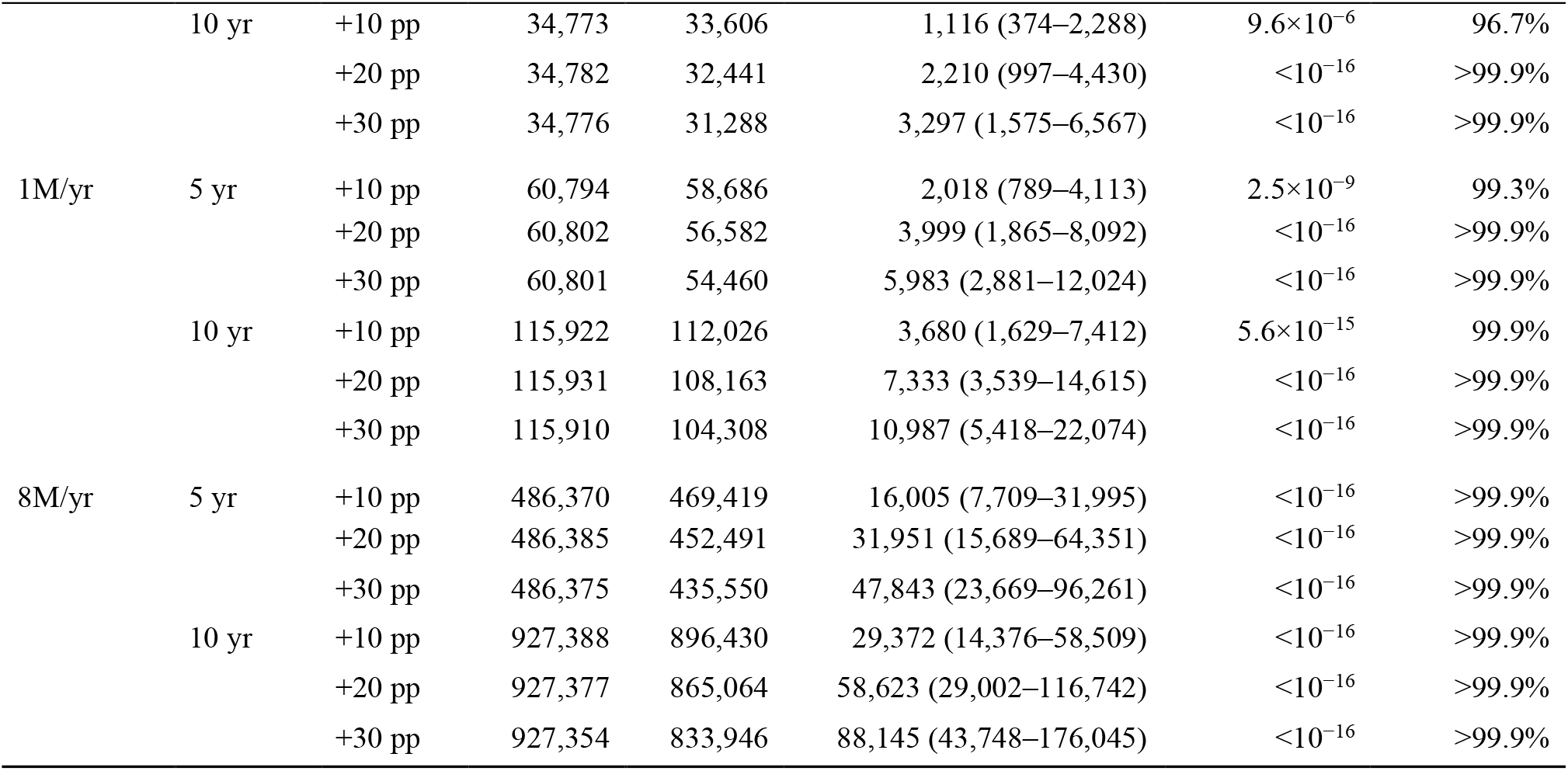
Full two-arm simulation results for all 18 design cells: median realized event totals in the control (*E*_ctrl_) and ROR (*E*_ror_) arms, the median between-arm difference *D*_*E*_ with its 95% uncertainty interval, the median one-sided *p*-value, and the assurance (expected power) at α = 0.05.

