## Supplementary material for "Projected Population-Level Impact of Digital Return of Results for Cardiovascular–Kidney–Metabolic Screening at US Blood Donation Centers: A Monte Carlo Simulation Study": Simulation Scripts: 00_model_functions.R

```
# SOAR CKM ROR Monte Carlo -- model functions
# Extracted verbatim (modulo file I/O) from Reports/SOAR_MC_Simulation_Official_V2.qmd
# so that all published numbers reproduce the V2 report exactly (seed 42, M = 100,000).

suppressPackageStartupMessages({
  library(dplyr); library(tidyr); library(tibble)
  library(purrr); library(extraDistr)
})

RANDOM_SEED  <- 42L
N_ITERATIONS <- 100000L
MU_B         <- 0.18     # control-arm action rate; used only by the two-arm check
KAPPA        <- 100

scenarios <- tibble::tibble(
  scenario = c("Carter BloodCare (300K/yr)",
               "Large Regional Center (1M/yr)",
               "National U.S. Pool (8M/yr)"),
  n_donors = c(300000, 1000000, 8000000)
)
time_horizons <- c(5L, 10L)
pp_scenarios  <- c(10L, 20L, 30L)   # ROR action-rate gain, percentage points

# Mean-preserving log-normal: E[draw] = mean exactly.
lognormal_draw <- function(mean, n, cv = 0.20) {
  sigma_sq <- log(1 + cv^2)
  mu       <- log(mean) - 0.5 * sigma_sq
  rlnorm(n, meanlog = mu, sdlog = sqrt(sigma_sq))
}

# Shared parameter draws (common random numbers across all design cells).
# The baseline action rate is drawn LAST, after every quantity the forecast
# uses, so that changing mu_b cannot perturb any other draw.
make_shared_draws <- function(n_iter, mu_b = MU_B, kappa = KAPPA) {
  stages <- extraDistr::rdirichlet(n_iter, c(55, 40, 5))
  tibble::tibble(
    stage0 = stages[, 1], stage1 = stages[, 2], stage2 = stages[, 3],
    mace_rrr = rbeta(n_iter, 30, 70),        # mean 0.30
    esrd_rrr = rbeta(n_iter, 40, 60),        # mean 0.40
    t2dm_rrr = rbeta(n_iter, 34, 66),        # mean 0.34
    mace_rate_s1 = lognormal_draw(0.005, n_iter),
    mace_rate_s2 = lognormal_draw(0.012, n_iter),
    esrd_rate_s1 = lognormal_draw(0.001, n_iter),
    esrd_rate_s2 = lognormal_draw(0.003, n_iter),
    t2dm_rate_s1 = lognormal_draw(0.021, n_iter),
    t2dm_rate_s2 = lognormal_draw(0.042, n_iter),
    baseline_rate = rbeta(n_iter, kappa * mu_b, kappa * (1 - mu_b))
  )
}

summarise_outcome <- function(v) {
  q <- quantile(v, probs = c(0.025, 0.25, 0.75, 0.975), names = FALSE, type = 7)
  tibble::tibble(
    median = as.integer(median(v)),
    lo95 = as.integer(q[1]), hi95 = as.integer(q[4]),
    lo50 = as.integer(q[2]), hi50 = as.integer(q[3]),
    mean = as.numeric(mean(v))
  )
}

# Intervention effect: strictly positive, E[Delta] = delta exactly.
lognormal_effect <- function(delta, n, cv = 0.30) {
  s2 <- log(1 + cv^2)
  rlnorm(n, meanlog = log(delta) - 0.5 * s2, sdlog = sqrt(s2))
}

# Per-actor events prevented: cumulative-incidence difference between the
# control hazard r and the treated hazard r(1 - rho) over `years`.
dci <- function(r, rho, years) (1 - r * (1 - rho))^years - (1 - r)^years

simulate_combo <- function(scenario, n_donors, years, pp, draws, n_iter) {
  Delta     <- lognormal_effect(pp / 100, n_iter)
  actors_s1 <- trunc(draws$stage1 * n_donors) * Delta
  actors_s2 <- trunc(draws$stage2 * n_donors) * Delta
  mace <- actors_s1 * dci(draws$mace_rate_s1, draws$mace_rrr, years) +
          actors_s2 * dci(draws$mace_rate_s2, draws$mace_rrr, years)
  esrd <- actors_s1 * dci(draws$esrd_rate_s1, draws$esrd_rrr, years) +
          actors_s2 * dci(draws$esrd_rate_s2, draws$esrd_rrr, years)
  t2dm <- actors_s1 * dci(draws$t2dm_rate_s1, draws$t2dm_rrr, years) +
          actors_s2 * dci(draws$t2dm_rate_s2, draws$t2dm_rrr, years)
  total <- mace + esrd + t2dm
  outcomes <- list(mace = mace, esrd = esrd, t2dm = t2dm, total = total)
  purrr::imap_dfr(outcomes, function(v, nm) {
    summarise_outcome(v) %>% tibble::add_column(outcome = nm, .before = 1)
  }) %>%
    tibble::add_column(scenario = scenario, n_donors = n_donors,
                       years = years, pp = pp, .before = 1)
}

run_all <- function(scenarios, time_horizons, pp_scenarios, draws, n_iter) {
  grid <- tidyr::expand_grid(
    scenario = scenarios$scenario, years = time_horizons, pp = pp_scenarios
  ) %>% dplyr::left_join(scenarios, by = "scenario")
  purrr::pmap_dfr(grid, function(scenario, years, pp, n_donors) {
    simulate_combo(scenario, n_donors, years, pp, draws, n_iter)
  })
}

# ---- Two-arm (control vs. ROR) trial simulation -----------------------------

cuminc <- function(r, years) 1 - (1 - r)^years

arm_events <- function(a, n1, n2, draws, years, n_iter) {
  so <- function(n_s, rate, rrr) {
    actors <- rbinom(n_iter, n_s, a)
    rbinom(n_iter, actors, cuminc(rate * (1 - rrr), years)) +
      rbinom(n_iter, n_s - actors, cuminc(rate, years))
  }
  so(n1, draws$mace_rate_s1, draws$mace_rrr) + so(n2, draws$mace_rate_s2, draws$mace_rrr) +
    so(n1, draws$esrd_rate_s1, draws$esrd_rrr) + so(n2, draws$esrd_rate_s2, draws$esrd_rrr) +
    so(n1, draws$t2dm_rate_s1, draws$t2dm_rrr) + so(n2, draws$t2dm_rate_s2, draws$t2dm_rrr)
}

simulate_trial <- function(scenario, n_donors, years, pp, draws, n_iter,
                           return_draws = FALSE) {
  B     <- draws$baseline_rate
  Delta <- lognormal_effect(pp / 100, n_iter)
  I     <- pmin(B + Delta, 1)
  n1    <- trunc(draws$stage1 * n_donors)
  n2    <- trunc(draws$stage2 * n_donors)
  E_ctrl    <- arm_events(B, n1, n2, draws, years, n_iter)
  E_ror     <- arm_events(I, n1, n2, draws, years, n_iter)
  prevented <- E_ctrl - E_ror
  z  <- prevented / sqrt(E_ctrl + E_ror)
  p1 <- pnorm(z, lower.tail = FALSE)
  # Full per-replication vectors (for figures); the RNG stream is identical
  # either way, so summary outputs are unaffected by this flag.
  if (return_draws) {
    return(tibble::tibble(E_ctrl = E_ctrl, E_ror = E_ror,
                          prevented = prevented, z = z, p1 = p1))
  }
  tibble::tibble(
    scenario = scenario, n_donors = n_donors, years = years, pp = pp,
    ctrl_med = as.integer(median(E_ctrl)), ror_med = as.integer(median(E_ror)),
    prev_med = as.integer(median(prevented)),
    prev_lo  = as.integer(quantile(prevented, 0.025, names = FALSE)),
    prev_hi  = as.integer(quantile(prevented, 0.975, names = FALSE)),
    pr_pos   = mean(prevented > 0), power = mean(p1 < 0.05),
    z_med    = median(z), p_med = median(p1)
  )
}
```
