## Supplementary material for "Projected Population-Level Impact of Digital Return of Results for Cardiovascular–Kidney–Metabolic Screening at US Blood Donation Centers: A Monte Carlo Simulation Study": Simulation Scripts: 01_run_forecast.R

```
# SOAR CKM ROR Monte Carlo -- forecast model (Section: Methods / Results)
# Reproduces the V2 report exactly: seed 42, M = 100,000, 18 design cells.
# Output: results/forecast_results.csv

args <- commandArgs(trailingOnly = FALSE)
this_dir <- dirname(sub("^--file=", "", args[grepl("^--file=", args)]))
source(file.path(this_dir, "00_model_functions.R"))
out_dir <- file.path(this_dir, "..", "results")
dir.create(out_dir, showWarnings = FALSE)

# Seeding sequence matches the V2 report: shared draws are generated first,
# then the RNG is re-seeded before the per-cell effect draws.
set.seed(RANDOM_SEED)
draws <- make_shared_draws(N_ITERATIONS)

set.seed(RANDOM_SEED)
res <- run_all(scenarios, time_horizons, pp_scenarios, draws, N_ITERATIONS)

write.csv(res, file.path(out_dir, "forecast_results.csv"), row.names = FALSE)
cat("Wrote", file.path(out_dir, "forecast_results.csv"), "-", nrow(res), "rows\n")

# Console check against the V2 reference cell (300K donors, 10 yr, +20 pp):
ref <- subset(res, n_donors == 3e5 & years == 10 & pp == 20 & outcome == "total")
cat(sprintf("Reference cell total: median %d, 95%% UI %d-%d (V2: 2201, 1099-4364)\n",
            ref$median, ref$lo95, ref$hi95))
```
