## Supplementary material for "Projected Population-Level Impact of Digital Return of Results for Cardiovascular–Kidney–Metabolic Screening at US Blood Donation Centers: A Monte Carlo Simulation Study": Simulation Scripts: 02_run_two_arm.R

```
# SOAR CKM ROR Monte Carlo -- two-arm (control vs. ROR) trial simulation
# Reproduces the V2 report exactly: same shared draws (seed 42), fresh seed
# before the trial loop, M = 100,000 simulated trials per design cell.
# Output: results/two_arm_results.csv

args <- commandArgs(trailingOnly = FALSE)
this_dir <- dirname(sub("^--file=", "", args[grepl("^--file=", args)]))
source(file.path(this_dir, "00_model_functions.R"))
out_dir <- file.path(this_dir, "..", "results")
dir.create(out_dir, showWarnings = FALSE)

set.seed(RANDOM_SEED)
draws <- make_shared_draws(N_ITERATIONS)

grid_ab <- tidyr::expand_grid(scenario = scenarios$scenario, years = time_horizons,
                              pp = pp_scenarios) %>%
  dplyr::left_join(scenarios, by = "scenario")

set.seed(RANDOM_SEED)
res_ab <- purrr::pmap_dfr(grid_ab, function(scenario, years, pp, n_donors) {
  simulate_trial(scenario, n_donors, years, pp, draws, N_ITERATIONS)
})

write.csv(res_ab, file.path(out_dir, "two_arm_results.csv"), row.names = FALSE)
cat("Wrote", file.path(out_dir, "two_arm_results.csv"), "-", nrow(res_ab), "rows\n")

ref <- subset(res_ab, n_donors == 3e5 & years == 10 & pp == 20)
cat(sprintf("Reference cell: D_E median %d, power %.4f, median p %.2e\n",
            ref$prev_med, ref$power, ref$p_med))
```
