## Supplementary material for "Projected Population-Level Impact of Digital Return of Results for Cardiovascular–Kidney–Metabolic Screening at US Blood Donation Centers: A Monte Carlo Simulation Study": Simulation Scripts: 03_tables_figures.R

```
# SOAR CKM ROR Monte Carlo -- manuscript tables and figures
# Reads results/*.csv and writes LaTeX table fragments and Figure 1 into results/.
# The manuscript pulls every number from these generated files (no hand-typing).

suppressPackageStartupMessages({ library(dplyr); library(ggplot2) })

args <- commandArgs(trailingOnly = FALSE)
this_dir <- dirname(sub("^--file=", "", args[grepl("^--file=", args)]))
res_dir  <- normalizePath(file.path(this_dir, "..", "results"))

res <- read.csv(file.path(res_dir, "forecast_results.csv"))
ab  <- read.csv(file.path(res_dir, "two_arm_results.csv"))

fmt  <- function(x) formatC(round(as.numeric(x)), format = "d", big.mark = ",")
val  <- function(sc, yr, p, oc, f = "median")
  res[res$n_donors == sc & res$years == yr & res$pp == p & res$outcome == oc, f]
shr  <- function(sc, yr, p, oc)
  100 * val(sc, yr, p, oc, "mean") / val(sc, yr, p, "total", "mean")

vols    <- c(3e5, 1e6, 8e6)
volslab <- c("300{,}000", "1{,}000{,}000", "8{,}000{,}000")
volsshort <- c("300K", "1M", "8M")
pps     <- c(10, 20, 30)
ppslab  <- c("$+10$ pp (conservative)", "$+20$ pp (primary)", "$+30$ pp (optimistic)")

writelines <- function(lines, file) {
  con <- file(file.path(res_dir, file)); writeLines(lines, con); close(con)
  cat("wrote", file, "\n")
}

## ---- Table 2 (main): total events prevented -------------------------------
lines <- c("\\begin{tabular}{llrrr}", "\\toprule",
  "Effect size & Horizon & \\multicolumn{1}{c}{300{,}000/yr} & \\multicolumn{1}{c}{1{,}000{,}000/yr} & \\multicolumn{1}{c}{8{,}000{,}000/yr} \\\\",
  "\\midrule")
for (j in seq_along(pps)) {
  p <- pps[j]
  if (j > 1) lines <- c(lines, "\\addlinespace")
  for (yr in c(5L, 10L)) {
    cells <- sapply(vols, function(k)
      sprintf("%s (%s--%s)", fmt(val(k, yr, p, "total")),
              fmt(val(k, yr, p, "total", "lo95")), fmt(val(k, yr, p, "total", "hi95"))))
    lab <- if (yr == 5L) ppslab[j] else ""
    lines <- c(lines, sprintf("%s & %d yr & %s \\\\", lab, yr, paste(cells, collapse = " & ")))
  }
}
lines <- c(lines, "\\bottomrule", "\\end{tabular}")
writelines(lines, "tab_events_main.tex")

## ---- Table 3 (main): NNS and screening cost per event ---------------------
lines <- c("\\begin{tabular}{lrrrr}", "\\toprule",
  " & \\multicolumn{2}{c}{5-year horizon} & \\multicolumn{2}{c}{10-year horizon} \\\\",
  "\\cmidrule(lr){2-3}\\cmidrule(lr){4-5}",
  "Effect size & \\multicolumn{1}{c}{NNS} & \\multicolumn{1}{c}{Cost per event} & \\multicolumn{1}{c}{NNS} & \\multicolumn{1}{c}{Cost per event} \\\\",
  "\\midrule")
for (j in seq_along(pps)) {
  p <- pps[j]
  cells <- unlist(lapply(c(5L, 10L), function(yr) {
    m <- val(3e5, yr, p, "total")
    c(fmt(3e5 / m), paste0("\\$", fmt(15 * 3e5 / m)))
  }))
  lines <- c(lines, sprintf("%s & %s \\\\", ppslab[j], paste(cells, collapse = " & ")))
}
lines <- c(lines, "\\bottomrule", "\\end{tabular}")
writelines(lines, "tab_nns_cost.tex")

## ---- Table 4 (main): assurance (expected power) of the two-arm design -----
vab <- function(sc, yr, p, f) ab[ab$n_donors == sc & ab$years == yr & ab$pp == p, f]
pw  <- function(x) { v <- 100 * x; if (v >= 99.95) "$>$99.9" else sprintf("%.1f", v) }
lines <- c("\\begin{tabular}{llrrr}", "\\toprule",
  "Donor volume & Horizon & \\multicolumn{1}{c}{$+10$ pp} & \\multicolumn{1}{c}{$+20$ pp} & \\multicolumn{1}{c}{$+30$ pp} \\\\",
  "\\midrule")
for (i in seq_along(vols)) {
  for (yr in c(5L, 10L)) {
    cells <- sapply(pps, function(p) pw(vab(vols[i], yr, p, "power")))
    lab <- if (yr == 5L) sprintf("%s/yr", volslab[i]) else ""
    lines <- c(lines, sprintf("%s & %d yr & %s \\\\", lab, yr, paste(cells, collapse = " & ")))
  }
  if (i < length(vols)) lines <- c(lines, "\\addlinespace")
}
lines <- c(lines, "\\bottomrule", "\\end{tabular}")
writelines(lines, "tab_assurance.tex")

## ---- Table S1: per-actor cumulative-incidence difference (saturation) -----
dci_fn <- function(r, rho, years) (1 - r * (1 - rho))^years - (1 - r)^years
dtab <- data.frame(
  outcome = c("MACE, Stage 1", "MACE, Stage 2", "ESRD, Stage 1",
              "ESRD, Stage 2", "T2DM, Stage 1", "T2DM, Stage 2"),
  r   = c(0.005, 0.012, 0.001, 0.003, 0.021, 0.042),
  rho = c(0.30,  0.30,  0.40,  0.40,  0.34,  0.34))
dtab$d5  <- dci_fn(dtab$r, dtab$rho, 5)
dtab$d10 <- dci_fn(dtab$r, dtab$rho, 10)
lines <- c("\\begin{tabular}{lrrrrr}", "\\toprule",
  "Outcome, stage & \\multicolumn{1}{c}{$r$} & \\multicolumn{1}{c}{$\\rho$} & \\multicolumn{1}{c}{$\\Delta\\mathrm{CI}$ ($T=5$)} & \\multicolumn{1}{c}{$\\Delta\\mathrm{CI}$ ($T=10$)} & \\multicolumn{1}{c}{10-yr\\,/\\,5-yr} \\\\",
  "\\midrule")
for (i in seq_len(nrow(dtab)))
  lines <- c(lines, sprintf("%s & %.3f & %.2f & %.4f & %.4f & %.2f \\\\",
    dtab$outcome[i], dtab$r[i], dtab$rho[i], dtab$d5[i], dtab$d10[i], dtab$d10[i] / dtab$d5[i]))
lines <- c(lines, "\\bottomrule", "\\end{tabular}")
writelines(lines, "tab_supp_dci.tex")

## ---- Table S2: full totals (median, IQR, 95% UI) for all 18 cells ---------
lines <- c("\\begin{tabular}{lllrrr}", "\\toprule",
  "Donor volume & Horizon & Effect & \\multicolumn{1}{c}{Median} & \\multicolumn{1}{c}{IQR} & \\multicolumn{1}{c}{95\\% UI} \\\\",
  "\\midrule")
for (i in seq_along(vols)) {
  for (yr in c(5L, 10L)) for (j in seq_along(pps)) {
    p <- pps[j]
    lab  <- if (yr == 5L && j == 1) sprintf("%s/yr", volslab[i]) else ""
    hlab <- if (j == 1) sprintf("%d yr", yr) else ""
    lines <- c(lines, sprintf("%s & %s & $+%d$ pp & %s & %s--%s & %s--%s \\\\",
      lab, hlab, p, fmt(val(vols[i], yr, p, "total")),
      fmt(val(vols[i], yr, p, "total", "lo50")), fmt(val(vols[i], yr, p, "total", "hi50")),
      fmt(val(vols[i], yr, p, "total", "lo95")), fmt(val(vols[i], yr, p, "total", "hi95"))))
  }
  if (i < length(vols)) lines <- c(lines, "\\addlinespace")
}
lines <- c(lines, "\\bottomrule", "\\end{tabular}")
writelines(lines, "tab_supp_totals.tex")

## ---- Table S3: outcome-specific medians and shares (primary effect size) --
lines <- c("\\begin{tabular}{llrrrr}", "\\toprule",
  "Donor volume & Horizon & \\multicolumn{1}{c}{MACE} & \\multicolumn{1}{c}{ESRD} & \\multicolumn{1}{c}{T2DM} & \\multicolumn{1}{c}{Total} \\\\",
  "\\midrule")
for (i in seq_along(vols)) {
  for (yr in c(5L, 10L)) {
    cells <- sapply(c("mace", "esrd", "t2dm"), function(oc)
      sprintf("%s (%.1f\\%%)", fmt(val(vols[i], yr, 20, oc)), shr(vols[i], yr, 20, oc)))
    lab <- if (yr == 5L) sprintf("%s/yr", volslab[i]) else ""
    lines <- c(lines, sprintf("%s & %d yr & %s & %s \\\\",
      lab, yr, paste(cells, collapse = " & "), fmt(val(vols[i], yr, 20, "total"))))
  }
  if (i < length(vols)) lines <- c(lines, "\\addlinespace")
}
lines <- c(lines, "\\bottomrule", "\\end{tabular}")
writelines(lines, "tab_supp_outcomes.tex")

## ---- Table S4: full two-arm results ---------------------------------------
pfmt <- function(p) {
  if (p >= 0.05) return(sprintf("$%.3f$", p))
  if (p < 1e-16) return("$<\\!10^{-16}$")
  e <- floor(log10(p)); m <- p / 10^e
  if (m >= 9.95) { m <- m / 10; e <- e + 1 }
  sprintf("$%.1f\\!\\times\\!10^{%d}$", m, e)
}
lines <- c("\\begin{tabular}{lllrrrrr}", "\\toprule",
  "Volume & Horizon & Effect & \\multicolumn{1}{c}{$E_{\\mathrm{ctrl}}$} & \\multicolumn{1}{c}{$E_{\\mathrm{ror}}$} & \\multicolumn{1}{c}{$D_E$ (95\\% UI)} & \\multicolumn{1}{c}{Median $p$} & \\multicolumn{1}{c}{Assurance} \\\\",
  "\\midrule")
for (i in seq_along(vols)) {
  for (yr in c(5L, 10L)) for (j in seq_along(pps)) {
    p <- pps[j]
    lab  <- if (yr == 5L && j == 1) sprintf("%s/yr", volsshort[i]) else ""
    hlab <- if (j == 1) sprintf("%d yr", yr) else ""
    lines <- c(lines, sprintf("%s & %s & $+%d$ pp & %s & %s & %s (%s--%s) & %s & %s\\%% \\\\",
      lab, hlab, p,
      fmt(vab(vols[i], yr, p, "ctrl_med")), fmt(vab(vols[i], yr, p, "ror_med")),
      fmt(vab(vols[i], yr, p, "prev_med")),
      fmt(vab(vols[i], yr, p, "prev_lo")), fmt(vab(vols[i], yr, p, "prev_hi")),
      pfmt(vab(vols[i], yr, p, "p_med")), pw(vab(vols[i], yr, p, "power"))))
  }
  if (i < length(vols)) lines <- c(lines, "\\addlinespace")
}
lines <- c(lines, "\\bottomrule", "\\end{tabular}")
writelines(lines, "tab_supp_two_arm.tex")

## ---- Figure 1: events prevented across the design grid --------------------
tot <- res %>% filter(outcome == "total") %>%
  mutate(
    vol = factor(n_donors, levels = rev(vols),
                 labels = rev(c("300,000/yr", "1,000,000/yr", "8,000,000/yr"))),
    eff = factor(pp, levels = pps, labels = c("+10 pp", "+20 pp", "+30 pp")),
    horizon = factor(years, levels = c(5, 10), labels = c("5-year horizon", "10-year horizon")))

pal <- c("+10 pp" = "#77AAD4", "+20 pp" = "#2E6FAC", "+30 pp" = "#123F6D")

p <- ggplot(tot, aes(y = vol, x = median, colour = eff, group = eff)) +
  geom_linerange(aes(xmin = lo95, xmax = hi95),
                 position = position_dodge(width = 0.62), linewidth = 0.45) +
  geom_linerange(aes(xmin = lo50, xmax = hi50),
                 position = position_dodge(width = 0.62), linewidth = 1.4) +
  geom_point(position = position_dodge(width = 0.62), size = 1.9) +
  facet_wrap(~horizon, ncol = 2) +
  scale_x_log10(breaks = c(300, 1000, 3000, 10000, 30000, 100000),
                labels = function(x) formatC(x, format = "d", big.mark = ",")) +
  scale_colour_manual(values = pal, name = "Action-rate gain",
                      guide = guide_legend(reverse = TRUE)) +
  labs(x = "ROR-attributable events prevented (log scale)", y = NULL) +
  theme_minimal(base_size = 9.5) +
  theme(panel.grid.minor = element_blank(),
        panel.grid.major.y = element_blank(),
        panel.grid.major.x = element_line(linewidth = 0.25, colour = "grey85"),
        axis.ticks.x = element_line(linewidth = 0.25, colour = "grey60"),
        strip.text = element_text(face = "bold", size = 9.5),
        legend.position = "bottom",
        legend.title = element_text(size = 9),
        plot.margin = margin(4, 8, 4, 4))

ggsave(file.path(res_dir, "fig2_events_prevented.pdf"), p,
       width = 6.8, height = 3.4, device = "pdf", useDingbats = FALSE)
cat("wrote fig2_events_prevented.pdf\n")
```
