## Supplementary material for "Projected Population-Level Impact of Digital Return of Results for Cardiovascular–Kidney–Metabolic Screening at US Blood Donation Centers: A Monte Carlo Simulation Study": Simulation Scripts: 04_two_arm_figures.R

```
# SOAR CKM ROR Monte Carlo -- Figure 3: control vs. ROR arm comparison
# Re-runs the full 18-cell two-arm loop with the identical RNG stream as
# 02_run_two_arm.R and captures the per-replication draws of the reference
# cell (300K donors, 10-year horizon, +20 pp), so the figure is exactly
# consistent with Table 4 / Table S4.
# Output: results/fig3_two_arm_distributions.pdf

suppressPackageStartupMessages({ library(dplyr); library(ggplot2); library(patchwork) })

args <- commandArgs(trailingOnly = FALSE)
this_dir <- dirname(sub("^--file=", "", args[grepl("^--file=", args)]))
source(file.path(this_dir, "00_model_functions.R"))
res_dir <- normalizePath(file.path(this_dir, "..", "results"))

set.seed(RANDOM_SEED)
draws <- make_shared_draws(N_ITERATIONS)

grid_ab <- tidyr::expand_grid(scenario = scenarios$scenario, years = time_horizons,
                              pp = pp_scenarios) %>%
  dplyr::left_join(scenarios, by = "scenario")

set.seed(RANDOM_SEED)
ref <- NULL
for (i in seq_len(nrow(grid_ab))) {
  g <- grid_ab[i, ]
  is_ref <- g$n_donors == 3e5 && g$years == 10L && g$pp == 20L
  out <- simulate_trial(g$scenario, g$n_donors, g$years, g$pp, draws,
                        N_ITERATIONS, return_draws = is_ref)
  if (is_ref) ref <- out
}

pr_pos <- mean(ref$prevented > 0)
cat(sprintf("Reference cell: median E_ctrl %d, E_ror %d, D_E %d; Pr(D_E>0) = %.5f\n",
            median(ref$E_ctrl), median(ref$E_ror), median(ref$prevented), pr_pos))

col_ctrl <- "#6E7B8B"   # neutral slate for the control arm
col_ror  <- "#2E6FAC"   # primary blue for the ROR arm (matches Figs 1-2)

arm_df <- rbind(
  data.frame(arm = "Control", events = ref$E_ctrl),
  data.frame(arm = "ROR",     events = ref$E_ror)
)

med_df <- arm_df %>% group_by(arm) %>% summarise(m = median(events), .groups = "drop")

base_thm <- theme_minimal(base_size = 9.5) +
  theme(panel.grid.minor = element_blank(),
        panel.grid.major.y = element_blank(),
        panel.grid.major.x = element_line(linewidth = 0.25, colour = "grey85"),
        axis.ticks.x = element_line(linewidth = 0.25, colour = "grey60"),
        legend.position = "none",
        plot.title = element_text(face = "bold", size = 9.5),
        plot.title.position = "plot")

# Display window: trim the extreme right tail so the central mass fills the
# panel and the between-arm shift is visible.
xwin <- quantile(arm_df$events, c(0.001, 0.99))
dctrl <- density(ref$E_ctrl, adjust = 1.1)
dror  <- density(ref$E_ror,  adjust = 1.1)
ymax  <- max(dctrl$y, dror$y)
m_ror  <- med_df$m[med_df$arm == "ROR"]
m_ctrl <- med_df$m[med_df$arm == "Control"]

pA <- ggplot(arm_df, aes(events, colour = arm, fill = arm)) +
  geom_density(alpha = 0.25, linewidth = 0.7, adjust = 1.1) +
  geom_vline(data = med_df, aes(xintercept = m, colour = arm),
             linewidth = 0.5, linetype = "22", show.legend = FALSE) +
  annotate("text", x = m_ror - 5500, y = ymax * 0.88,
           label = sprintf("ROR\nmedian %s", format(m_ror, big.mark = ",")),
           colour = col_ror, size = 2.9, fontface = "bold",
           hjust = 1, vjust = 1, lineheight = 1.05) +
  annotate("text", x = m_ctrl + 7500, y = ymax * 0.80,
           label = sprintf("Control\nmedian %s", format(m_ctrl, big.mark = ",")),
           colour = col_ctrl, size = 2.9, fontface = "bold",
           hjust = 0, vjust = 1, lineheight = 1.05) +
  scale_colour_manual(values = c(Control = col_ctrl, ROR = col_ror)) +
  scale_fill_manual(values = c(Control = col_ctrl, ROR = col_ror)) +
  scale_x_continuous(labels = function(x) format(x, big.mark = ",", scientific = FALSE),
                     breaks = seq(20000, 60000, 10000)) +
  scale_y_continuous(expand = expansion(mult = c(0, 0.12))) +
  coord_cartesian(xlim = xwin) +
  labs(title = "A  Realized event totals by arm",
       x = "Events over 10 years", y = "Density") +
  base_thm +
  theme(axis.text.y = element_blank(), panel.grid.major.x = element_blank())

q <- quantile(ref$prevented, c(0.025, 0.975))
dens <- density(ref$prevented, adjust = 1.5)
dd <- data.frame(x = dens$x, y = dens$y)

pB <- ggplot(dd, aes(x, y)) +
  geom_area(data = subset(dd, x >= q[1] & x <= q[2]),
            fill = "#2E6FAC", alpha = 0.25) +
  geom_line(colour = "#2E6FAC", linewidth = 0.7) +
  geom_vline(xintercept = 0, linewidth = 0.5, colour = "grey30", linetype = "22") +
  geom_vline(xintercept = median(ref$prevented),
             colour = "#123F6D", linewidth = 0.5) +
  annotate("text", x = median(ref$prevented) + 150, y = max(dd$y) * 0.95,
           label = sprintf("median %s", format(median(ref$prevented), big.mark = ",")),
           colour = "#123F6D", size = 3.0, hjust = 0) +
  annotate("text", x = 150, y = max(dd$y) * 0.35, label = "0",
           colour = "grey30", size = 3.0, hjust = 0) +
  scale_x_continuous(labels = function(x) format(x, big.mark = ",", scientific = FALSE)) +
  scale_y_continuous(expand = expansion(mult = c(0, 0.05))) +
  labs(title = "B  Between-arm difference (events prevented)",
       x = expression("Difference"~italic(D)[E]~"over 10 years"),
       y = "Density") +
  base_thm +
  theme(axis.text.y = element_blank(), panel.grid.major.x = element_blank())

p <- pA + pB

ggsave(file.path(res_dir, "fig3_two_arm_distributions.pdf"), p,
       width = 6.8, height = 2.7, device = "pdf", useDingbats = FALSE)
cat("wrote fig3_two_arm_distributions.pdf\n")

writeLines(sprintf("pr_pos_reference_cell,%.6f", pr_pos),
           file.path(res_dir, "two_arm_reference_stats.csv"))
```
