## Supplementary material for "Projected Population-Level Impact of Digital Return of Results for Cardiovascular–Kidney–Metabolic Screening at US Blood Donation Centers: A Monte Carlo Simulation Study": Simulation Scripts: 05_assurance_curves.R

```
# SOAR CKM ROR Monte Carlo -- Figure 4: assurance as a function of the
# action-rate gain. Evaluates the two-arm design over a finer effect grid
# (+2 to +30 pp in 2-pp steps) at every volume and horizon, using the same
# shared parameter draws (seed 42, M = 100,000) as the main analysis.
# Outputs: results/assurance_curve.csv, results/fig4_assurance_curves.pdf

suppressPackageStartupMessages({ library(dplyr); library(ggplot2) })

args <- commandArgs(trailingOnly = FALSE)
this_dir <- dirname(sub("^--file=", "", args[grepl("^--file=", args)]))
source(file.path(this_dir, "00_model_functions.R"))
res_dir <- normalizePath(file.path(this_dir, "..", "results"))

set.seed(RANDOM_SEED)
draws <- make_shared_draws(N_ITERATIONS)

pp_fine <- seq(2L, 30L, by = 2L)
grid_fine <- tidyr::expand_grid(scenario = scenarios$scenario, years = time_horizons,
                                pp = pp_fine) %>%
  dplyr::left_join(scenarios, by = "scenario")

set.seed(RANDOM_SEED)
res_fine <- purrr::pmap_dfr(grid_fine, function(scenario, years, pp, n_donors) {
  simulate_trial(scenario, n_donors, years, pp, draws, N_ITERATIONS)
})

write.csv(res_fine, file.path(res_dir, "assurance_curve.csv"), row.names = FALSE)
cat("wrote assurance_curve.csv -", nrow(res_fine), "rows\n")

# Smallest gain reaching 80% / 90% assurance, per volume and horizon.
for (thr in c(0.80, 0.90)) {
  cat(sprintf("-- assurance >= %.0f%%:\n", 100 * thr))
  th <- res_fine %>% group_by(n_donors, years) %>% arrange(pp) %>%
    summarise(min_pp = if (any(power >= thr)) min(pp[power >= thr]) else NA_integer_,
              .groups = "drop")
  print(as.data.frame(th))
}

pal <- c("300,000/yr" = "#77AAD4", "1,000,000/yr" = "#2E6FAC",
         "8,000,000/yr" = "#123F6D")

plot_df <- res_fine %>%
  mutate(vol = factor(n_donors, levels = c(3e5, 1e6, 8e6),
                      labels = names(pal)),
         horizon = factor(years, levels = c(5, 10),
                          labels = c("5-year horizon", "10-year horizon")))

p <- ggplot(plot_df, aes(pp, 100 * power, colour = vol, group = vol)) +
  geom_hline(yintercept = c(80, 90), linewidth = 0.35,
             linetype = "22", colour = "grey60") +
  geom_line(linewidth = 0.8) +
  geom_point(size = 1.4) +
  facet_wrap(~horizon, ncol = 2) +
  scale_x_continuous(breaks = seq(0, 30, 5),
                     labels = function(x) sprintf("+%d", x)) +
  scale_y_continuous(limits = c(0, 101), breaks = seq(0, 100, 20),
                     expand = expansion(mult = c(0, 0.01))) +
  scale_colour_manual(values = pal, name = "Annual donor volume") +
  annotate("text", x = 30, y = 76.5, label = "80%", colour = "grey45",
           size = 2.7, hjust = 1) +
  annotate("text", x = 30, y = 93.5, label = "90%", colour = "grey45",
           size = 2.7, hjust = 1) +
  labs(x = "Action-rate gain (percentage points)", y = "Assurance (%)") +
  theme_minimal(base_size = 9.5) +
  theme(panel.grid.minor = element_blank(),
        panel.grid.major.x = element_blank(),
        panel.grid.major.y = element_line(linewidth = 0.25, colour = "grey85"),
        axis.ticks.x = element_line(linewidth = 0.25, colour = "grey60"),
        strip.text = element_text(face = "bold", size = 9.5),
        legend.position = "bottom",
        legend.title = element_text(size = 9))

ggsave(file.path(res_dir, "fig4_assurance_curves.pdf"), p,
       width = 6.8, height = 3.2, device = "pdf", useDingbats = FALSE)
cat("wrote fig4_assurance_curves.pdf\n")
```
